## supplemental material for "Hypoxaemia prevalence and management among children and adults presenting to primary care facilities in Uganda: a prospective cohort study"

**FIGURE: MAP OF UGANDA**
**Map of Uganda with study regions/districts highlighted**

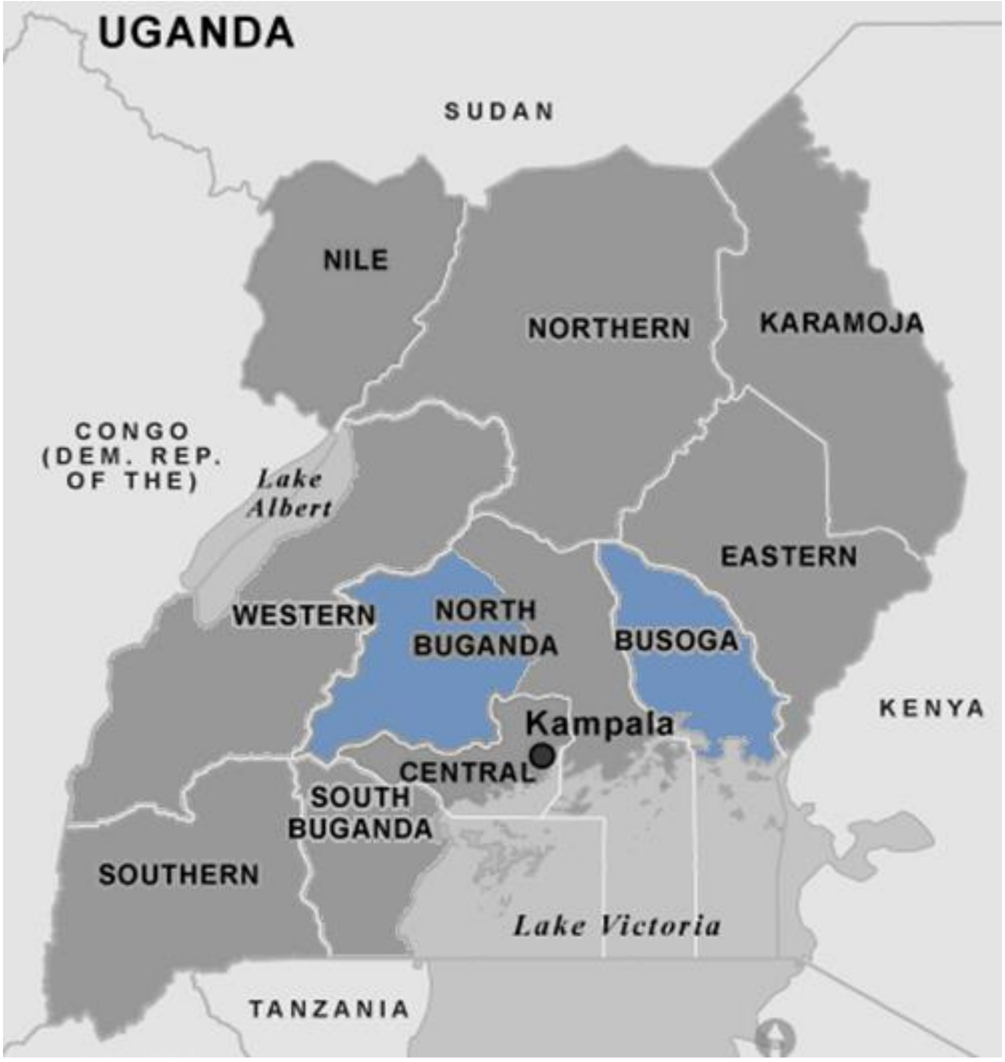

**TABLE: PARTICIPANT CHARACTERISTICS**

**Demographic and clinical features of 5780 acutely unwell children, adolescents, and adults presenting to primary care (HCIII) facilities in Uganda, Feb-Apr 2021**

| **DEMOGRAPHIC INFORMATION** | | | | | | | |
|---|---|---|---|---|---|---|---|
| **Age group** | Busoga | | North Central | | Total | | |
| Neonate | 10 | 0.2% | 6 | 0.4% | 16 | 0.3% | |
| 1-11 mths | 280 | 6.8% | 96 | 5.8% | 376 | 6.5% | |
| 1-4 yrs | 911 | 22.1% | *258* | 15.5% | 1169 | 20.2% | |
| 5-9yrs | 469 | 11.4% | *164* | 9.9% | 633 | 11.0% | |
| 10-14 yrs | 202 | 4.9% | *100* | 6.0% | 302 | 5.2% | |
| 15-24 yrs | 793 | 19.2% | *295* | 17.8% | 1088 | 18.8% | |
| 25-49 yrs | 1,072 | 26.0% | 530 | 31.9% | 1602 | 27.7% | |
| 50+ yrs | 383 | 9.3% | 211 | 12.7% | 594 | 10.3% | |
| *Total* | *4120* | | *1660* | | *5780* | | |
| *<5 years* | 1201 | 29.2% | 360 | 21.7% | *1561* | 27.0% | |
| *5-14 yrs* | 671 | 16.3% | 264 | 15.9% | *935* | 16.2% | |
| *>= 15yrs* | 2248 | 54.6% | 1036 | 62.4% | *3284* | 56.8% | *P<0.001* |
| **Sex** | Busoga | | North Central | | Total | | |
| Male | 1207 | 29.3% | 654 | 39.4% | 1861 | 32.2% | |
| Female | 2913 | 70.7% | 1006 | 60.6% | 3919 | 67.8% | *P<0.001* |
| **Vital signs documented** | Busoga | | North Central | | Total | | |
| Temperature | 303 | 7.4% | 159 | 9.6% | 462 | 8.0% | *P=0.005* |
| Heart rate | 2867 | 69.6% | 1036 | 62.4% | 3903 | 67.5% | *P<0.001* |
| Respiratory rate | 12 | 0.3% | 2 | 0.1% | 14 | 0.2% | *P=0.232* |
| Blood pressure | 98 | 2.4% | 57 | 3.4% | 155 | 2.7% | *P=0.024* |
| **PRESENTING COMPLAINTS** | | | | | | | |
| **Fever/chills** | Busoga | | North Central | | Total | | |
| *<5 yrs* | 1,002 | 83.4% | 206 | 57.2% | 1208 | 77.4% | |
| *5-14 yrs* | 481 | 71.7% | 146 | 55.3% | 627 | 67.1% | |
| >=15 yrs | 871 | 38.7% | 459 | 44.3% | 1330 | 40.5% | |
| *Total* | *2354* | *57.1%* | *811* | *48.9%* | *3165* | *54.8%* | |
| **Respiratory complaints** | Busoga | | North Central | | Total | | |
| *<5 yrs* | 885 | 73.7% | 273 | 75.8% | 1158 | 74.2% | |
| *5-14 yrs* | 367 | 54.7% | 144 | 54.5% | 511 | 54.7% | |
| >=15 yrs | 800 | 35.6% | 400 | 38.6% | 1200 | 36.5% | |
| *Total* | *2052* | *49.8%* | *817* | *49.2%* | *2869* | *49.6%* | |
| **Fever/chills OR Respiratory** | Busoga | | North Central | | Total | | |
| *<5 yrs* | 1,148 | 95.6% | 330 | 91.7% | 1478 | 94.7% | |
| *5-14 yrs* | 585 | 87.2% | 208 | 78.8% | 793 | 84.8% | |
| >=15 yrs | 1329 | 59.1% | 686 | 66.2% | 2015 | 61.4% | |
| *Total* | *3062* | *74.3%* | *1224* | *73.7%* | *4286* | *74.2%* | |
| **Pain** | Busoga | | North Central | | Total | | |
| *<5 yrs* | 147 | 12.2% | 37 | 10.3% | 184 | 11.8% | |
| *5-14 yrs* | 359 | 53.5% | 89 | 33.7% | 448 | 47.9% | |
| >=15 yrs | 1611 | 71.7% | 554 | 53.5% | 2165 | 65.9% | |
| *Total* | *2117* | *51.4%* | *680* | *41.0%* | *2797* | *48.4%* | |
| **Abdominal complaints** | Busoga | | North Central | | Total | | |
| *<5 yrs* | 233 | 19.4% | 39 | 10.8% | 272 | 17.4% | |
| *5-14 yrs* | 258 | 38.5% | 77 | 29.2% | 335 | 35.8% | |
| *>= 15yrs* | 905 | 40.3% | 344 | 33.2% | 1249 | 38.0% | |

| | Busoga | | North Central | | Total | |
|---|---|---|---|---|---|---|
| *Total* | *1396* | *33.9%* | *460* | *27.7%* | *1856* | *32.1%* |
| **Diarrhoea or vomiting** | Busoga | | North Central | | Total | |
| *<5 yrs* | 362 | 30.1% | 97 | 26.9% | 459 | 29.4% |
| *5-14 yrs* | 131 | 19.5% | 41 | 15.5% | 172 | 18.4% |
| >=15 yrs | 153 | 6.8% | 79 | 7.6% | 232 | 7.1% |
| *Total* | *646* | *15.7%* | *217* | *13.1%* | *863* | *14.9%* |
| **Urogenital complaints** | Busoga | | North Central | | Total | |
| *<5 yrs* | 10 | 0.8% | 5 | 1.4% | 15 | 1.0% |
| *5-14 yrs* | 15 | 2.2% | 7 | 2.7% | 22 | 2.4% |
| >=15 yrs | 208 | 9.3% | 103 | 9.9% | 311 | 9.5% |
| *Total* | *233* | *5.7%* | *115* | *6.9%* | *348* | *6.0%* |
| **Skin complaints** | Busoga | | North Central | | Total | |
| *<5 yrs* | 54 | 4.5% | 21 | 5.8% | 75 | 4.8% |
| *5-14 yrs* | 21 | 3.1% | 11 | 4.2% | 32 | 3.4% |
| >=15 yrs | 39 | 1.7% | 30 | 2.9% | 69 | 2.1% |
| *Total* | *114* | *2.8%* | *62* | *3.7%* | *176* | *3.0%* |
| **Ear, eye, mouth complaints** | Busoga | | North Central | | Total | |
| *<5 yrs* | 27 | 2.2% | 25 | 6.9% | 52 | 3.3% |
| *5-14 yrs* | 17 | 2.5% | 18 | 6.8% | 35 | 3.7% |
| >=15 yrs | 48 | 2.1% | 34 | 3.3% | 82 | 2.5% |
| *Total* | *92* | *2.2%* | *77* | *4.6%* | *169* | *2.9%* |
| **All other complaints** | Busoga | | North Central | | Total | |
| *<5 yrs* | 277 | 23.1% | 51 | 14.2% | 328 | 21.0% |
| *5-14 yrs* | 137 | 20.4% | 38 | 14.4% | 175 | 18.7% |
| >=15 yrs | 623 | 27.7% | 207 | 20.0% | 830 | 25.3% |
| *Total* | *1037* | *25.2%* | *296* | *17.8%* | *1333* | *23.1%* |
| **Other complaint as only complaint** | Busoga | | North Central | | Total | |
| *<5 yrs* | 8 | 0.7% | 6 | 1.7% | 14 | 0.9% |
| *5-14 yrs* | 2 | 0.3% | 6 | 2.3% | 8 | 0.9% |
| >=15 yrs | 34 | 1.5% | 29 | 2.8% | 63 | 1.9% |
| *Total* | *44* | *1.1%* | *41* | *2.5%* | *85* | *1.5%* |
| **DIAGNOSES** | | | | | | |
| **Malaria** | Busoga | | North Central | | Total | |
| *<5 yrs* | 751 | 62.5% | 74 | 20.6% | 825 | 52.9% |
| *5-14 yrs* | 461 | 68.7% | 77 | 29.2% | 538 | 57.5% |
| >=15 yrs | 799 | 35.5% | 166 | 16.0% | 965 | 29.4% |
| *Total* | *2011* | *48.8%* | *317* | *19.1%* | *2328* | *40.3%* |
| **ARI** | Busoga | | North Central | | Total | |
| *<5 yrs* | 570 | 47.5% | 241 | 66.9% | 811 | 52.0% |
| *5-14 yrs* | 242 | 36.1% | 121 | 45.8% | 363 | 38.8% |
| >=15 yrs | 648 | 28.8% | 356 | 34.4% | 1004 | 30.6% |
| *Total* | *1460* | *35.4%* | *718* | *43.3%* | *2178* | *37.7%* |
| **Diarrhoeal disease** | Busoga | | North Central | | Total | |
| *<5 yrs* | 178 | 14.8% | 65 | 18.1% | 243 | 15.6% |
| *5-14 yrs* | 39 | 5.8% | 18 | 6.8% | 57 | 6.1% |
| >=15 yrs | 84 | 3.7% | 29 | 2.8% | 113 | 3.4% |
| *Total* | *301* | *7.3%* | *112* | *6.7%* | *413* | *7.1%* |
| **Helminths** | Busoga | | North Central | | Total | |
| *<5 yrs* | 27 | 2.2% | 14 | 3.9% | 41 | 2.6% |
| *5-14 yrs* | 28 | 4.2% | 18 | 6.8% | 46 | 4.9% |

| | | | | | | |
|---|---|---|---|---|---|---|
| >=15 yrs | 46 | 2.0% | 20 | 1.9% | 66 | 2.0% |
| *Total* | *101* | *2.5%* | *52* | *3.1%* | *153* | *2.6%* |
| **Skin disorders** | Busoga | | North Central | | Total | |
| *<5 yrs* | 43 | 3.6% | 26 | 7.2% | 69 | 4.4% |
| *5-14 yrs* | 16 | 2.4% | 11 | 4.2% | 27 | 2.9% |
| >=15 yrs | 43 | 1.9% | 20 | 1.9% | 63 | 1.9% |
| *Total* | *102* | *2.5%* | *57* | *3.4%* | *159* | *2.8%* |
| **STI/Genital/UTI** | Busoga | | North Central | | Total | |
| *<5 yrs* | 4 | 0.3% | 3 | 0.8% | 7 | 0.4% |
| *5-14 yrs* | 10 | 1.5% | 10 | 3.8% | 20 | 2.1% |
| >=15 yrs | 404 | 18.0% | 216 | 20.8% | 620 | 18.9% |
| *Total* | *418* | *10.1%* | *229* | *13.8%* | *647* | *11.2%* |
| **Septicaemia** | Busoga | | North Central | | Total | |
| *<5 yrs* | 34 | 2.8% | 7 | 1.9% | 41 | 2.6% |
| *5-14 yrs* | 9 | 1.3% | 5 | 1.9% | 14 | 1.5% |
| >=15 yrs | 36 | 1.6% | 20 | 1.9% | 56 | 1.7% |
| *Total* | *79* | *1.9%* | *32* | *1.9%* | *111* | *1.9%* |
| **Non-diarrhoeal digestive system illnesses** | Busoga | | North Central | | Total | |
| *<5 yrs* | 6 | 0.5% | 2 | 0.6% | 8 | 0.5% |
| *5-14 yrs* | 10 | 1.5% | 6 | 2.3% | 16 | 1.7% |
| >=15 yrs | 280 | 12.5% | 95 | 9.2% | 375 | 11.4% |
| *Total* | *296* | *7.2%* | *103* | *6.2%* | *399* | *6.9%* |

Notes: Presenting complaints as reported by patient/caregiver. Diagnosis as recorded by treating healthcare worker. P-values obtained from Pearson's chi-squared or Fishers exact test as applicable. ARI = acute respiratory infection; STI – sexually transmitted infection; UTI – urinary tract infection.

### TABLE: HYPOXAEMIA PREVALENCE (EXTENDED RESULTS)

**Prevalence of hypoxaemia among acutely unwell children, adolescents and adults presenting to HCIII facilities in Uganda, Feb-Apr 2021**

| | N | % | Severe hypoxaemia ($SpO_2$<90%) | | | | | Moderate hypoxaemia ($SpO_2$ 90-93%) | | | | |
|---|---|---|---|---|---|---|---|---|---|---|---|---|
| | | | N | % | prevalence | 95% CI | | N | % | prevalence | 95% CI | |
| **Neonate** | 16 | 0.3% | 0 | . | . | . | . | 4 | 3.9% | 25.0% | 8.9% | 53.3% |
| **1-11 mths** | 376 | 6.5% | 8 | 29.6% | 2.1% | 1.1% | 4.2% | 27 | 26.2% | 7.2% | 5.0% | 10.3% |
| **1-4 yrs** | 1,169 | 20.2% | 13 | 48.1% | 1.1% | 0.6% | 1.9% | 45 | 43.7% | 3.8% | 2.9% | 5.1% |
| **5-9yrs** | 633 | 11.0% | 3 | 11.1% | 0.5% | 0.2% | 1.5% | 5 | 4.9% | 0.8% | 0.3% | 1.9% |
| **10-14 yrs** | 302 | 5.2% | 0 | . | . | . | . | 5 | 4.9% | 1.7% | 0.7% | 3.9% |
| **15-24 yrs** | 1,088 | 18.8% | 0 | . | . | . | . | 3 | 2.9% | 0.3% | 0.1% | 0.9% |
| **25-49 yrs** | 1,602 | 27.7% | 1 | 3.7% | 0.1% | 0.0% | 0.4% | 7 | 6.8% | 0.4% | 0.2% | 0.9% |
| **50+ yrs** | 594 | 10.3% | 2 | 7.4% | 0.3% | 0.1% | 1.3% | 7 | 6.8% | 1.2% | 0.6% | 2.5% |
| ***Total*** | **5780** | **100%** | **27** | **100%** | **0.5%** | **0.3%** | **0.7%** | **103** | **100%** | **1.8%** | **1.5%** | **2.2%** |
| **Girls (<5 years)** | 790 | 13.7% | 14 | 51.9% | 1.8% | 1.1% | 3.0% | 38 | 36.9% | 4.8% | 3.5% | 6.5% |
| **Boys (<5 years)** | 771 | 13.3% | 7 | 25.9% | 0.9% | 0.4% | 1.9% | 38 | 36.9% | 4.9% | 3.6% | 6.7% |
| **Girls (5-14 years)** | 568 | 9.8% | 1 | 3.7% | 0.2% | 0.0% | 1.2% | 4 | 3.9% | 0.7% | 0.3% | 1.9% |
| **Boys (5-14 years)** | 367 | 6.3% | 2 | 7.4% | 0.5% | 0.1% | 2.2% | 6 | 5.8% | 1.6% | 0.7% | 3.6% |
| **Women (≥15 years)** | 2,561 | 44.3% | 1 | 3.7% | 0.0% | 0.0% | 0.3% | 14 | 13.6% | 0.5% | 0.3% | 0.9% |
| **Men (≥15 years)** | 723 | 12.5% | 2 | 7.4% | 0.3% | 0.1% | 1.1% | 3 | 2.9% | 0.4% | 0.1% | 1.3% |
| ***Total*** | **5,780** | **100%** | **27** | **100%** | **0.5%** | **0.3%** | **0.7%** | **103** | **100%** | **1.8%** | **1.5%** | **2.2%** |
| **Busoga** | 4,120 | 71.3% | 20 | 74.1% | 0.5% | 0.3% | 0.7% | 86 | 83.5% | 2.1% | 1.7% | 2.6% |
| **North Central** | 1,660 | 28.7% | 7 | 25.9% | 0.4% | 0.2% | 0.9% | 17 | 16.5% | 1.0% | 0.6% | 1.6% |
| ***Total*** | **5,780** | **100%** | **27** | **100%** | **0.5%** | **0.3%** | **0.7%** | **103** | **100%** | **1.8%** | **1.5%** | **2.2%** |

#### HYPOXAEMIA PREVALENCE BY PRESENTING COMPLAINT

| | N | % | Severe hypoxaemia ($SpO_2$<90%) | | | | | Moderate hypoxaemia ($SpO_2$ 90-93%) | | | | |
|---|---|---|---|---|---|---|---|---|---|---|---|---|
| | | | N | % | prevalence | 95% CI | | N | % | prevalence | 95% CI | |
| **U5 years by presenting complaints** | | | | | | | | | | | | |
| **Abdominal** | 272 | 17.4% | 2 | 9.5% | 0.7% | 0.2% | 2.9% | 5 | 6.6% | 1.8% | 0.8% | 4.4% |
| **Urogenital** | 15 | 1.0% | 0 | . | . | . | . | 3 | 3.9% | 20.0% | 5.9% | 50.0% |
| **Resp OR fever** | 1,478 | 94.7% | 21 | 100% | 1.4% | 0.9% | 2.2% | 75 | 98.7% | 5.1% | 4.1% | 6.3% |
| **Respiratory** | 1,158 | 74.2% | 18 | 85.7% | 1.6% | 1.0% | 2.5% | 67 | 88.2% | 5.8% | 4.6% | 7.3% |

| | | | | | | | | | | | | |
|---|---|---|---|---|---|---|---|---|---|---|---|---|
| **Fever** | 1,208 | 77.4% | 15 | 71.4% | 1.2% | 0.7% | 2.1% | 59 | 77.6% | 4.9% | 3.8% | 6.3% |
| **Diarrhoeal** | 459 | 29.4% | 5 | 23.8% | 1.1% | 0.5% | 2.6% | 17 | 22.4% | 3.7% | 2.3% | 5.9% |
| **Skin** | 75 | 4.8% | 0 | . | . | . | . | 1 | 1.3% | 1.3% | 0.2% | 9.1% |
| **Eye, ear, dental** | 52 | 3.3% | 1 | 4.8% | 1.9% | 0.3% | 13.0% | 0 | . | . | . | . |
| **Pain NOS** | 184 | 11.8% | 0 | . | . | . | . | 7 | 9.2% | 3.8% | 1.8% | 7.8% |
| **All other** | 328 | 21.0% | 11 | 52.4% | 3.4% | 1.9% | 6.0% | 25 | 32.9% | 7.6% | 5.2% | 11.1% |
| **Other unique** | 14 | 0.9% | 0 | . | . | . | . | 0 | . | . | . | . |
| ***Total <5yrs*** | **1,561** | ***** | **21** | ***** | **1.3%** | **0.9%** | **2.1%** | **76** | ***** | **4.9%** | **3.9%** | **6.1%** |
| **5-14 years by presenting complaints** | | | | | | | | | | | | |
| **Abdominal** | 335 | 35.8% | 0 | . | . | . | . | 2 | 20.0% | 0.6% | 0.1% | 2.4% |
| **Urogenital** | 22 | 2.4% | 0 | . | . | . | . | 0 | . | . | . | . |
| **Resp OR fever** | 793 | 84.8% | 3 | 100% | 0.4% | 0.1% | 1.2% | 10 | 100% | 1.3% | 0.7% | 2.3% |
| **Respiratory** | 511 | 54.7% | 3 | 100% | 0.6% | 0.2% | 1.8% | 8 | 80.0% | 1.6% | 0.8% | 3.1% |
| **Fever** | 627 | 67.1% | 3 | 100% | 0.5% | 0.2% | 1.5% | 8 | 80.0% | 1.3% | 0.6% | 2.5% |
| **Diarrhoeal** | 172 | 18.4% | 0 | . | . | . | . | 2 | 20.0% | 1.2% | 0.3% | 4.6% |
| **Skin** | 32 | 3.4% | 0 | . | . | . | . | 0 | . | . | . | . |
| **Eye, ear, dental** | 35 | 3.7% | 0 | . | . | . | . | 0 | . | . | . | . |
| **Pain NOS** | 448 | 47.9% | 3 | 100% | 0.7% | 0.2% | 2.1% | 4 | 40.0% | 0.9% | 0.3% | 2.4% |
| **All other** | 175 | 18.7% | 1 | 33.3% | 0.6% | 0.1% | 4.0% | 4 | 40.0% | 2.3% | 0.9% | 6.0% |
| **Other unique** | 8 | 0.9% | 0 | . | . | . | . | 0 | . | . | . | . |
| ***Total 5-14 years*** | **935** | ***** | **3** | ***** | **0.3%** | **0.1%** | **1.0%** | **10** | ***** | **1.1%** | **0.6%** | **2.0%** |
| **15+ years by presenting complaints** | | | | | | | | | | | | |
| **Abdominal** | 1,249 | 38.0% | 0 | . | . | . | . | 4 | 23.5% | 0.3% | 0.1% | 0.9% |
| **Urogenital** | 311 | 9.5% | 0 | . | . | . | . | 1 | 5.9% | 0.3% | 0.0% | 2.3% |
| **Resp OR fever** | 2,015 | 61.4% | 2 | 66.7% | 0.1% | 0.0% | 0.4% | 11 | 64.7% | 0.5% | 0.3% | 1.0% |
| **Respiratory** | 1,200 | 36.5% | 2 | 66.7% | 0.2% | 0.0% | 0.7% | 7 | 41.2% | 0.6% | 0.3% | 1.2% |
| **Fever** | 1,330 | 40.5% | 1 | 33.3% | 0.1% | 0.0% | 0.5% | 6 | 35.3% | 0.5% | 0.2% | 1.0% |
| **Diarrhoeal** | 232 | 7.1% | 0 | . | . | . | . | 1 | 5.9% | 0.4% | 0.1% | 3.0% |
| **Skin** | 69 | 2.1% | 0 | . | . | . | . | 2 | 11.8% | 2.9% | 0.7% | 11.1% |
| **Eye, ear, dental** | 82 | 2.5% | 0 | . | . | . | . | 0 | . | . | . | . |
| **Pain NOS** | 2,165 | 65.9% | 2 | 66.7% | 0.1% | 0.0% | 0.4% | 11 | 64.7% | 0.5% | 0.3% | 0.9% |
| **All other** | 830 | 25.3% | 2 | 66.7% | 0.2% | 0.1% | 1.0% | 7 | 41.2% | 0.8% | 0.4% | 1.8% |
| **Other unique** | 63 | 1.9% | 0 | . | . | . | . | 1 | 5.9% | 1.6% | 0.2% | 10.8% |

| | | | | | | | | | | | | |
|---|---|---|---|---|---|---|---|---|---|---|---|---|
| ***Total cases <15yrs*** | **3,284** | ***** | **3** | ***** | **0.1%** | **0.0%** | **0.3%** | **17** | ***** | **0.5%** | **0.3%** | **0.8%** |

**HYPOXAEMIA PREVALENCE BY CLINICIAN DIAGNOSIS**

| | N | % | Severe hypoxaemia ($SpO_2$<90%) | | | | | Moderate hypoxaemia ($SpO_2$ 90-93%) | | | | |
|---|---|---|---|---|---|---|---|---|---|---|---|---|
| | | | N | % | | | | | N | % | | |
| **U5 years by diagnosis** | | | | | | | | | | | | |
| **ARI** | 811 | 52.0% | 13 | 61.9% | 1.6% | 0.9% | 2.7% | 53 | 69.7% | 6.5% | 5.0% | 8.5% |
| **ARI (no pneumonia)** | 678 | 43.4% | 6 | 28.6% | 0.9% | 0.4% | 2.0% | 35 | 46.1% | 5.2% | 3.7% | 7.1% |
| **"Pneumonia"** | 133 | 8.5% | 7 | 33.3% | 5.3% | 2.5% | 10.7% | 18 | 23.7% | 13.5% | 8.7% | 20.5% |
| **Diarrhoeal disease** | 243 | 15.6% | 3 | 14.3% | 1.2% | 0.4% | 3.8% | 10 | 13.2% | 4.1% | 2.2% | 7.5% |
| **Other GIT** | 8 | 0.5% | 0 | . | . | . | . | 0 | . | . | . | . |
| **Allergies** | 6 | 0.4% | 0 | . | . | . | . | 0 | . | . | . | . |
| **Ear infections** | 12 | 0.8% | 0 | . | . | . | . | 1 | 1.3% | 8.3% | 0.9% | 47.5% |
| **Injury** | 8 | 0.5% | 1 | 4.8% | 12.5% | 1.1% | 64.2% | 0 | . | . | . | . |
| **STI/Genital/UTI** | 7 | 0.4% | 0 | . | . | . | . | 1 | 1.3% | 14.3% | 1.2% | 70.1% |
| **Malaria** | 825 | 52.9% | 10 | 47.6% | 1.2% | 0.7% | 2.2% | 34 | 44.7% | 4.1% | 3.0% | 5.7% |
| **Sepsis** | 41 | 2.6% | 0 | . | . | . | . | 7 | 9.2% | 17.1% | 8.2% | 32.3% |
| **Helminths** | 41 | 2.6% | 0 | . | . | . | . | 1 | 1.3% | 2.4% | 0.3% | 16.2% |
| **Pregnancy** | 0 | . | . | . | . | . | . | | . | . | . | . |
| **Skin** | 69 | 4.4% | 1 | 4.8% | 1.4% | 0.2% | 9.9% | 1 | 1.3% | 1.4% | 0.2% | 9.9% |
| **All other Dx** | 67 | 4.3% | 1 | 4.8% | 1.5% | 0.2% | 10.2% | 4 | 5.3% | 6.0% | 2.2% | 15.1% |
| **Other unique** | 23 | 1.5% | 0 | . | . | . | . | 0 | . | . | . | . |
| ***Total <5yrs*** | **1,561** | ***** | **21** | ***** | **1.3%** | **0.9%** | **2.1%** | **76** | ***** | **4.9%** | **3.9%** | **6.1%** |
| **5-14 years by diagnosis** | | | | | | | | | | | | |
| **ARI** | 363 | 38.8% | 2 | 66.7% | 0.6% | 0.1% | 2.2% | 4 | 40.0% | 1.1% | 0.4% | 2.9% |
| **ARI (no pneumonia)** | 354 | 37.9% | 1 | 33.3% | 0.3% | 0.0% | 2.0% | 4 | 40.0% | 1.1% | 0.4% | 3.0% |
| **"Pneumonia"** | 9 | 1.0% | 1 | 33.3% | 11.1% | 1.1% | 59.1% | 0 | . | . | . | . |
| **Diarrhoeal disease** | 57 | 6.1% | 0 | . | . | . | . | 1 | 10.0% | 1.8% | 0.2% | 11.9% |
| **Other GIT** | 16 | 1.7% | 0 | . | . | . | . | 0 | . | . | . | . |
| **Allergies** | 7 | 0.7% | 0 | . | . | . | . | 0 | . | . | . | . |
| **Ear infections** | 10 | 1.1% | 0 | . | . | . | . | 0 | . | . | . | . |
| **Injury** | 6 | 0.6% | 0 | . | . | . | . | 0 | . | . | | . |

| | | | | | | | | | | | | |
|---|---|---|---|---|---|---|---|---|---|---|---|---|
| **STI/Genital/UTI** | 20 | 2.1% | 0 | . | . | . | . | 0 | . | . | . | . |
| **Malaria** | 538 | 57.5% | 1 | 33.3% | 0.2% | 0.0% | 1.3% | 5 | 50.0% | 0.9% | 0.4% | 2.2% |
| **Sepsis** | 14 | 1.5% | 0 | . | . | . | . | 0 | . | . | . | . |
| **Helminths** | 46 | 4.9% | 0 | . | . | . | . | 0 | . | . | . | . |
| **Pregnancy** | 0 | . | . | . | . | . | . | . | . | . | . | . |
| **Skin** | 27 | 2.9% | 0 | . | . | . | . | 0 | . | . | . | . |
| **All other Dx** | 59 | 6.3% | 2 | 66.7% | 3.4% | 0.8% | 12.9% | 2 | 20.0% | 3.4% | 0.8% | 12.9% |
| **Other unique** | 37 | 4.0% | 1 | 33.3% | 2.7% | 0.4% | 17.8% | 1 | 10.0% | 2.7% | 0.4% | 17.8% |
| ***Total 5-14yrs*** | **935** | ***** | **3** | ***** | **0.3%** | **0.1%** | **1.0%** | **10** | | **1.1%** | **0.6%** | **2.0%** |
| **15+ years by diagnosis** | | | | | | | | | | | | |
| **ARI** | 1,004 | 30.6% | 0 | . | . | . | . | 6 | 35.3% | 0.6% | 0.3% | 1.3% |
| **ARI (no pneumonia)** | 990 | 30.1% | 0 | . | . | . | . | 6 | 35.3% | 0.6% | 0.3% | 1.3% |
| **"Pneumonia"** | 14 | 0.4% | 0 | . | . | . | . | 0 | . | . | . | . |
| **Diarrhoeal disease** | 113 | 3.4% | 0 | . | . | . | . | 0 | . | . | . | . |
| **Other GIT** | 375 | 11.4% | 0 | . | . | . | . | 2 | 11.8% | 0.5% | 0.1% | 2.1% |
| **Allergies** | 30 | 0.9% | 0 | . | . | . | . | 1 | 5.9% | 3.3% | 0.4% | 21.6% |
| **Ear infections** | 23 | 0.7% | 0 | . | . | . | . | 0 | . | . | . | . |
| **Injury** | 48 | 1.5% | 0 | . | . | . | . | 0 | . | . | . | . |
| **STI/Genital/UTI** | 620 | 18.9% | 0 | . | . | . | . | 3 | 17.6% | 0.5% | 0.2% | 1.5% |
| **Malaria** | 965 | 29.4% | 1 | 33.3% | 0.1% | 0.0% | 0.7% | 3 | 17.6% | 0.3% | 0.1% | 1.0% |
| **Sepsis** | 56 | 1.7% | 0 | . | . | . | . | 0 | . | . | . | . |
| **Helminths** | 66 | 2.0% | 0 | . | . | . | . | 1 | 5.9% | 1.5% | 0.2% | 10.3% |
| **Pregnancy** | 41 | 1.2% | 0 | . | . | . | . | 0 | . | . | . | . |
| **Skin** | 63 | 1.9% | 0 | . | . | . | . | 1 | 5.9% | 1.6% | 0.2% | 10.8% |
| **All other Dx** | 588 | 17.9% | 1 | 33.3% | 0.2% | 0.0% | 1.2% | 4 | 23.5% | 0.7% | 0.3% | 1.8% |
| **Other unique** | 373 | 11.4% | 1 | 33.3% | 0.3% | 0.0% | 1.9% | 2 | 11.8% | 0.5% | 0.1% | 2.1% |
| ***Total 15+yrs*** | **3,284** | ***** | **3** | | **0.1%** | **0.0%** | **0.3%** | **17** | ***** | **0.5%** | **0.3%** | **0.8%** |

**FIGURE: HYPOXAEMIA PREVALENCE ACROSS AGE GROUPS**

**Bimodal age distribution of hypoxaemia ($SpO_2$<94%) among acutely unwell children, adolescents, and adults presenting to HCIII facilities in Uganda, Feb-Apr 2021.**

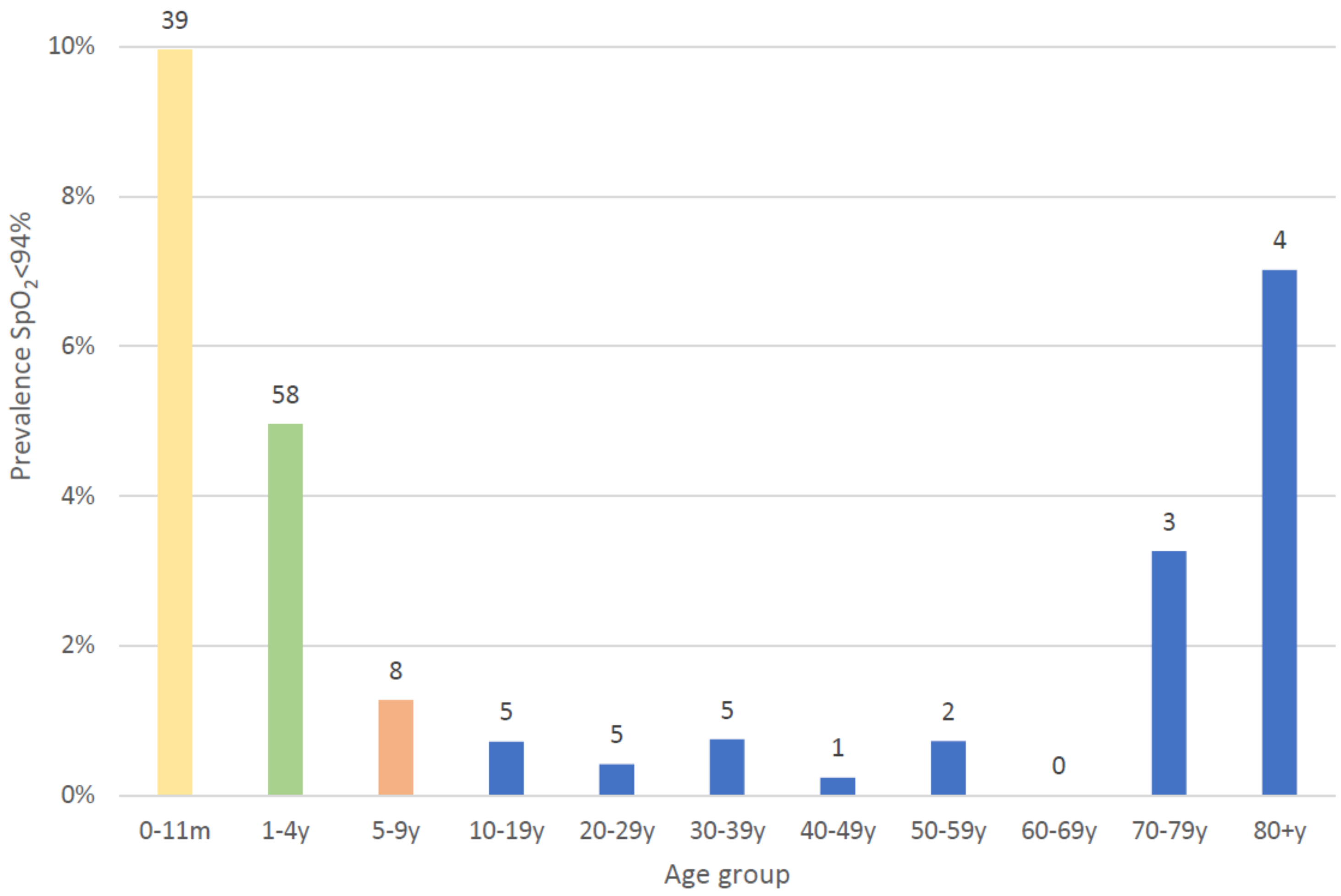

Notes: Number labels on each bar represent the number of hypoxaemia cases.

### TABLE: PREDICTORS OF HYPOXAEMIA

**Predictors of hypoxaemia among children, adolescents, and adults presenting to HCIII facilities in Uganda, using mixed-effects logistic regression**

| ***Final model*** | **Under 5** | | | | **Under 15** | | | | **5-14 years** | | | | **15+** | | | |
|---|---|---|---|---|---|---|---|---|---|---|---|---|---|---|---|---|
| | **aOR** | **95%** | **CI** | **p** | **aOR** | **95%** | **CI** | **p** | **aOR** | **95%** | **CI** | **p** | **aOR** | **95%** | **CI** | **p** |
| **Age** | 0.66 | 0.54 | 0.79 | 0.00 | 0.79 | 0.73 | 0.86 | 0.00 | . | . | | . | 1.05 | 1.02 | 1.07 | 0.00 |
| **Respiratory complaints** | 2.46 | 1.30 | 4.66 | 0.01 | 2.85 | 1.59 | 5.12 | 0.00 | 5.11 | 1.10 | 23.73 | 0.04 | . | . | | . |
| | **ICC** | **95%** | **CI** | | **ICC** | **95%** | **CI** | | **ICC** | **95%** | **CI** | | **ICC** | **95%** | **CI** | |
| **Facility** | 0.21 | 0.09 | 0.42 | | 0.18 | 0.08 | 0.36 | | 0.18 | 0.02 | 0.69 | | 0.31 | 0.09 | 0.67 | |

| ***Full model*** | **Under 5 years** | | | | **Under 15 years** | | | | **5-14 years** | | | | **15+ years** | | | |
|---|---|---|---|---|---|---|---|---|---|---|---|---|---|---|---|---|
| | **aOR** | **95%** | **CI** | **p** | **aOR** | **95%** | **CI** | **p** | **aOR** | **95%** | **CI** | **p** | **aOR** | **95%** | **CI** | **p** |
| **Region** | 0.43 | 0.17 | 1.09 | 0.08 | 0.60 | 0.26 | 1.37 | 0.22 | 1.82 | 0.41 | 8.00 | 0.43 | 0.58 | 0.13 | 2.60 | 0.47 |
| **Age (years)** | 0.67 | 0.55 | 0.81 | 0.00 | 0.80 | 0.73 | 0.88 | 0.00 | 1.10 | 0.89 | 1.37 | 0.38 | 1.05 | 1.02 | 1.07 | 0.00 |
| **Sex** | 1.22 | 0.79 | 1.88 | 0.36 | 1.05 | 0.70 | 1.56 | 0.82 | 0.42 | 0.13 | 1.35 | 0.15 | 1.27 | 0.43 | 3.80 | 0.66 |
| **Abdominal complaints** | 0.61 | 0.27 | 1.39 | 0.24 | 0.52 | 0.25 | 1.07 | 0.08 | 0.51 | 0.11 | 2.49 | 0.41 | 0.46 | 0.14 | 1.49 | 0.20 |
| **Urogenital complaints** | . | . | | . | . | . | | . | . | . | | . | 0.65 | 0.08 | 5.32 | 0.69 |
| **Respiratory complaints** | 2.24 | 1.16 | 4.30 | 0.02 | 2.53 | 1.39 | 4.61 | 0.00 | 5.43 | 1.10 | 26.85 | 0.04 | 1.13 | 0.44 | 2.92 | 0.80 |
| **Diarrhoea and vomiting** | 0.75 | 0.44 | 1.25 | 0.27 | 0.79 | 0.49 | 1.29 | 0.35 | 1.36 | 0.27 | 6.96 | 0.71 | 0.90 | 0.11 | 7.18 | 0.92 |
| **Fever and chills** | 0.87 | 0.51 | 1.48 | 0.60 | 1.01 | 0.61 | 1.66 | 0.97 | 3.20 | 0.66 | 15.59 | 0.15 | 0.74 | 0.28 | 1.98 | 0.55 |
| **Pain** | 0.83 | 0.36 | 1.93 | 0.67 | 0.95 | 0.50 | 1.79 | 0.86 | 1.84 | 0.56 | 5.99 | 0.31 | 0.60 | 0.21 | 1.69 | 0.33 |
| **Intercept** | 0.67 | 0.02 | 18.15 | 0.81 | 0.19 | 0.01 | 3.54 | 0.26 | 0.00 | 0.00 | 0.11 | 0.01 | 0.00 | 0.00 | 1.44 | 0.07 |
| | **ICC** | **95%** | **CI** | | **ICC** | **95%** | **CI** | | **ICC** | **95%** | **CI** | | **ICC** | **95%** | **CI** | |
| **Facility** | 0.18 | 0.07 | 0.39 | | 0.15 | 0.06 | 0.34 | | 0.22 | 0.22 | 0.22 | | 0.29 | 0.08 | 0.66 | |

Notes: Final model = after backward stepwise selection; CI = confidence interval; ICC – inter-cluster correlation coefficient;

### TABLE: PREDICTORS OF REFERRAL

**Predictors of referral among children, adolescents, and adults presenting to HCIII facilities in Uganda, using mixed-effects logistic regression**

| *Final model* | **Under 5 years** | | | | **Under 15 years** | | | | **5-14 years** | | | | **15+ years** | | | |
|---|---|---|---|---|---|---|---|---|---|---|---|---|---|---|---|---|
| | **aOR** | **95%** | **CI** | **p** | **aOR** | **95%** | **CI** | **p** | **aOR** | **95%** | **CI** | **p** | **aOR** | **95%** | **CI** | **p** |
| **Region** | 18.5 | 1.03 | 331.8 | 0.05 | | | | | | | | | 19.5 | 3.69 | 102.5 | 0.00 |
| **Age (years)** | 0.24 | 0.07 | 0.88 | 0.03 | | | | | | | | | 1.03 | 1.00 | 1.05 | 0.03 |
| **Sex** | | | | | | | | | | | | | | | | |
| **$SpO_2$<94%** | 960 | 30.7 | 29973 | 0.00 | 41.0 | 13.9 | 121.2 | 0.00 | 19.1 | 1.98 | 183.6 | 0.01 | 58.0 | 9.95 | 338.6 | 0.00 |
| **Abdominal complaints** | 16.0 | 1.22 | 211.1 | 0.04 | | | | | | | | | | | | |
| **Urogenital complaints** | . | . | | . | | | | | | | | | | | | |
| **Respiratory complaints** | 0.06 | 0.01 | 0.61 | 0.02 | | | | | | | | | | | | |
| **Diarrhoea and vomiting** | | | | | | | | | | | | | | | | |
| **Skin complaints** | | | | | | | | | | | | | | | | |
| **Fever and chills** | | | | | | | | | | | | | | | | |
| **Pain NOS** | | | | | | | | | | | | | | | | |
| | **ICC** | **95%** | **CI** | | **ICC** | **95%** | **CI** | | **ICC** | **95%** | **CI** | | **ICC** | **95%** | **CI** | |
| **Facility** | 0.47 | 0.13 | 0.84 | | 0.17 | 0.03 | 0.57 | | NA | | | | 0.09 | 0.01 | 0.54 | |

Notes: Final model = after backward stepwise selection; CI = confidence interval; ICC – inter-cluster correlation coefficient;

**FIGURE: FLOW CHART FOR CHILDREN WITH SEVERE HYPOXAEMIA ($SpO_2$<90%)**

**Flow chart showing referral, facility attendance, and day 7 outcomes for 24 children with severe hypoxaemia SpO2<90%**

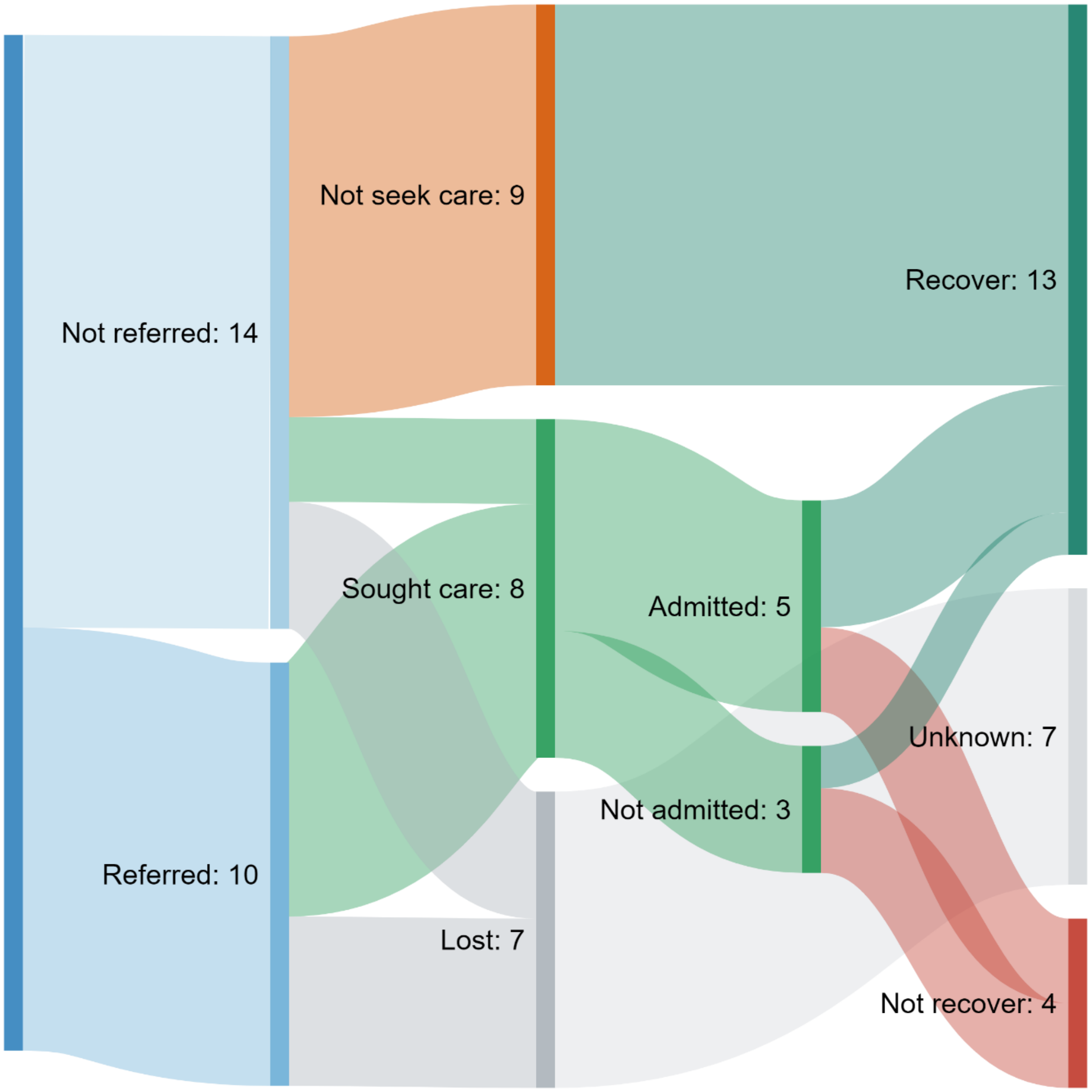
